# Metastatic vs. Localized Disease As Inclusion Criteria That Can Be Automatically Extracted From Randomized Controlled Trials Using Natural Language Processing

**DOI:** 10.1101/2024.06.17.24309020

**Authors:** Paul Windisch, Fabio Dennstädt, Carole Koechli, Robert Förster, Christina Schröder, Daniel M. Aebersold, Daniel R. Zwahlen

**Affiliations:** Department of Radiation Oncology, Cantonal Hospital Winterthur, Winterthur, Switzerland; Department of Radiation Oncology, Inselspital, Bern University Hospital, University of Bern, Bern, Switzerland

**Author notes:** **Correspondence:** Paul Windisch, MD, Department of Radiation Oncology, Kantonsspital Winterthur, Brauerstrasse 15, Haus R, 8400 Winterthur. **Ethics approval and consent to participate:** Not applicable. **Availability of data and materials:** All data and code used to obtain this study’s results have been uploaded to https://github.com/windisch-paul/metastatic_vs_local. **Funding:** No funding was received for this project. **Author contributions:** Conceptualization, P.W., F.D., C.K.; methodology, P.W, D.R.Z.; formal analysis, P.W.; data curation, P.W.; writing—original draft preparation, P.W.; writing—review and editing, F.D., C.K., R.F., C.S., D.M.A., D.R.Z.; supervision, D.R.Z.; project administration, D.M.A., D.R.Z. All authors read and approved the final manuscript.

**Keywords:** Natural language processing, Randomized controlled trial, Evidence-based medicine, Metastases, Machine Learning, Transformer

## Abstract

**Background:** Extracting inclusion and exclusion criteria in a structured, automated fashion remains a challenge to developing better search functionalities or automating systematic reviews of randomized controlled trials in oncology. The question “Did this trial enroll patients with localized disease, metastatic disease, or both?” could be used to narrow down the number of potentially relevant trials when conducting a search.

**Methods:** 600 trials from high-impact medical journals were classified depending on whether they allowed for the inclusion of patients with localized and/or metastatic disease. 500 trials were used to develop and validate three different models with 100 trials being stored away for testing.

**Results:** On the test set, a rule-based system using regular expressions achieved an F1-score of 0.72 (95% CI: 0.64 - 0.81) for the prediction of whether the trial allowed for the inclusion of patients with localized disease and 0.77 (95% CI: 0.69 - 0.85) for metastatic disease. A transformer-based machine learning model achieved F1 scores of 0.97 (95% CI: 0.93 - 1.00) and 0.88 (95% CI: 0.82 - 0.94), respectively. The best performance was achieved by a combined approach where the rule-based system was allowed to overrule the machine learning model with F1 scores of 0.97 (95% CI: 0.94 - 1.00) and 0.89 (95% CI: 0.83 - 0.95), respectively.

**Conclusion:** Automatic classification of cancer trials with regard to the inclusion of patients with localized and or metastatic disease is feasible. Turning the extraction of trial criteria into classification problems could, in selected cases, improve text-mining approaches in evidence-based medicine.

## Introduction

Automating the extraction of PICO (patient, intervention, control, outcome) characteristics from randomized controlled trials (RCTs) using natural language processing (NLP) could have various advantages, from assessing adherence to reporting standards over using metadata for filtering trials to automating systematic reviews and meta-analyses.^1,2^ This has created interest from both academic and commercial entities to improve the quality of reporting and to rapidly identify trials that provide information that is relevant to a particular clinical questions.^3^

Currently available tools, such as Trialstreamer, are mostly domain-agnostic and process RCTs from any field.^4^ When it comes to inclusion and exclusion criteria, they are often able to identify and extract the paragraphs that provide the information. However, ideally, the extraction would go even further and result in a structured list of criteria that either apply or don’t apply (e.g., Is a history of prior malignancy allowed? Yes/No).^5^

In oncology, a common eligibility criterion is the extent of the disease, which can be provided in various ways, including the TNM stage, keywords like advanced, extensive, or metastatic, and numerical thresholds, e.g., regarding the size of the primary tumor or the number of lymph nodes.^6^ Therefore, it is hard for models to extract this information in a structured fashion, as other factors, like the tumor entity, also influence the TNM stage.^7^

A key question that could be used to narrow down the number of potentially relevant trials in the absence of perfect inclusion criteria extraction could be: “Did this trial enroll patients with localized disease, metastatic disease, or both?” This precisely defined question has a set of allowable responses and can therefore be used as a common data element (CDE) in a variety of scenarios such as currently recruiting trials or manuscripts of published trials to ensure consistent data collection.^8^

Therefore, we trained a machine learning model to answer this question for published clinical trials based on their abstracts. As the feasibility of classifying abstracts according to different criteria has been demonstrated previously^9^, our hypothesis was that this classification task would result in sufficiently good performance to automatically annotate trials with this information and allow for filtering according to this criterion.

## Methods

For this prototype, randomized controlled oncology trials from seven major journals (British Medical Journal, JAMA, JAMA Oncology, Journal of Clinical Oncology, Lancet, Lancet Oncology, New England Journal of Medicine) published between 2005 and 2023 were randomly sampled and annotated with the labels “LOCAL”, “METASTATIC”, both or none. Trials that allowed for the inclusion of patients with localized disease received the label “LOCAL”. Trials that allowed for the inclusion of patients with metastatic disease received the label “METASTATIC”. Trials that allowed for the inclusion of patients with either localized or metastatic disease received bot labels. Screening trials that enrolled patients without known cancer or trials of interventions to prevent cancer were assigned no label. Trials of tumor entities where the distinction between localized and metastatic disease is usually not made (e.g., hematologic malignancies) were skipped. Annotation was based on the title and abstract. If those were inconclusive, the full text of the publication was evaluated. If this was not conclusive either, the registration or the protocol were evaluated. Annotation was performed by a single author (P.W.) using the tool prodigy (v. 1.13.1).

We hypothesized that the modeling should result in good performance with a limited number of examples as the phraseology that is used to report the information tends to repeat itself. Due to this and also in order to limit the workload for this prototype, annotation was stopped after 600 trials with the option of increasing the training data later on. Trials were then randomly shuffled, and 100 trials were stored away for testing.

Prior to training a machine learning (ML) model, we developed a rule-based system using regular expressions (i.e., expressions that match certain patterns in text) to take advantage of the repeating phraseology and to serve as our baseline. The rule-based system consisted of several conditions that checked the title of a publication to judge whether the trial allowed for the inclusion of patients with localized and/or metastatic disease (e.g., searching for the term “metastatic” but not “non-metastatic”, stages, etc.). If no condition applied, the system performed a random guess. The conditions were defined based on the training/validation set. The full list of regular expressions can be found in the repository for this article (https://github.com/windisch-paul/metastatic_vs_local).

The titles and abstracts of 500 trials were used to train and validate a multilabel text classification model using a random 80:20 split. The transformer model RoBERTa-base was trained using Adam as the optimizer.^10,11^ The detailed configuration file with all parameters that were used for training and validation is available from the repository.

Lastly, we also developed a combined approach that used the ML model’s output as its preliminary prediction, which could be overruled if one of the conditions triggered by the regular expressions applied.

All three models (rule-based, ML, and combined) were tested on the same previously unseen test set. A threshold of 0.5 was used to assign predictions to a class. 95% confidence intervals were estimated using normal approximation intervals. Training, validation, and testing were performed in python (version 3.11.5) using, among others, the pandas (version 2.1.0), spacy (version 3.7.4), and spacy-transformers (1.2.5) packages.

## Results

Figure 1 presents the distribution of trials that allowed for the inclusion of patients with localized and metastatic disease in the training/validation and test set. 71.6% of trials in the training/validation set and 77% of trials in the test set allowed for the inclusion of patients with localized disease. 61.2% of trials in the training/validation set and 51% of trials in the test set allowed for the inclusion of patients with metastatic disease.

**Figure 1.**
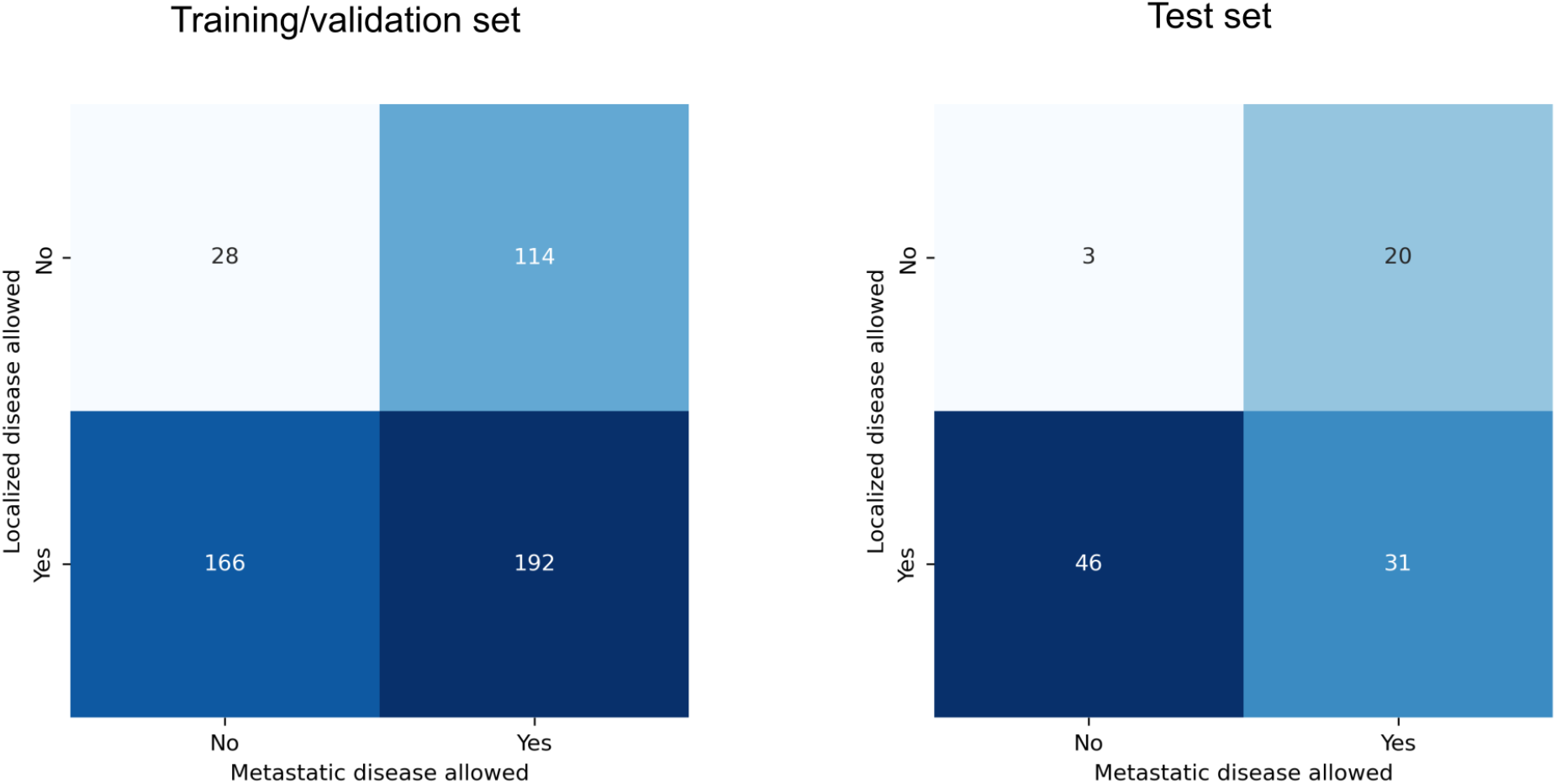
Distribution of trials that allowed for the inclusion of patients with localized and metastatic disease in the training/validation and test set.

The best-performing model during training achieved an F1 score of 0.89 on the validation set when predicting whether a trial allowed for the inclusion of patients with localized disease (precision 0.83, recall 0.96). For metastatic disease, the F1 score was also 0.89 (precision 0.81, recall 1.00).

All performance metrics on the test set, including confidence intervals, can be found in Table 1. The confusion matrices are presented in Figure 2.

**Table 1.** Performance metrics on the test set. Numbers in parentheses indicate the 95% confidence intervals; *ML = Machine Learning*.

|  |  | Accuracy | Precision | Recall | F1-Score |
| --- | --- | --- | --- | --- | --- |
| Rule-based | Localized disease | 0.61 (0.51 - 0.71) | 0.80 (0.70 - 0.90) | 0.66 (0.56 - 0.77) | 0.72 (0.64 - 0.81) |
|  | Metastatic disease | 0.73 (0.64 - 0.82) | 0.68 (0.57 - 0.79) | 0.88 (0.79 - 0.97) | 0.77 (0.69 - 0.85) |
| ML | Localized disease | 0.95 (0.91 - 0.99) | 0.95 (0.90 - 1.00) | 0.99 (0.96 - 1.00) | 0.97 (0.93 - 1.00) |
|  | Metastatic disease | 0.86 (0.79 - 0.93) | 0.78 (0.68 - 0.88) | 1.00 (1.00 - 1.00) | 0.88 (0.82 - 0.94) |
| Combined | Localized disease | 0.96 (0.92 - 1.00) | 0.95 (0.90 - 1.00) | 1.00 (1.00 - 1.00) | 0.97 (0.94 - 1.00) |
|  | Metastatic disease | 0.88 (0.83 - 0.95) | 0.81 (0.71 - 0.91) | 1.00 (1.00 - 1.00) | 0.89 (0.83 - 0.95) |

**Figure 2.**
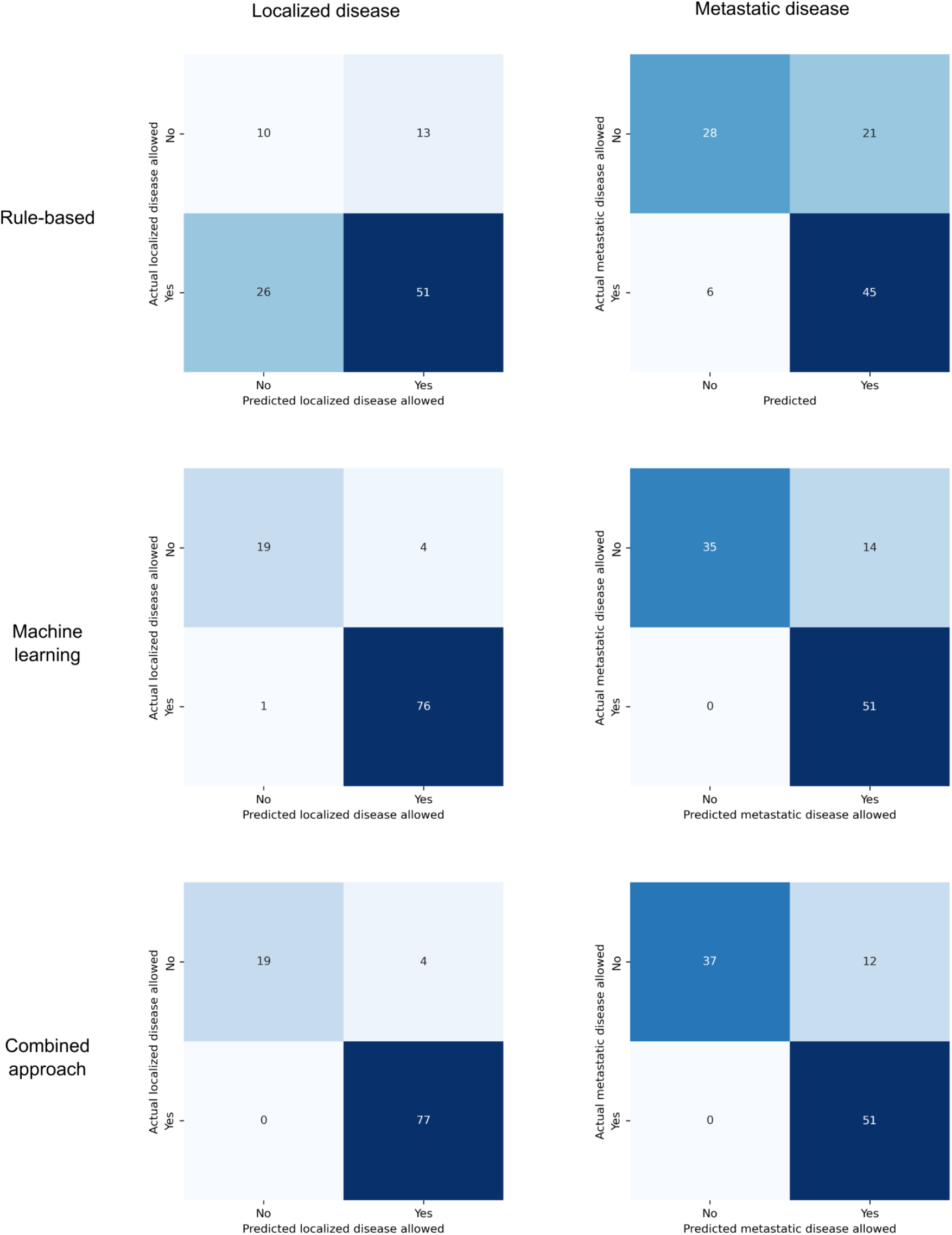
Confusion matrices for the performance of the rule-based system (top), the machine learning model (middle), and the combined approach of refining the machine learning model using rules (bottom) on predicting whether patients with localized disease (left) and/or metastatic disease (right) were eligible for a trial.

The rule-based model achieved accuracies of 0.61 and 0.73 for predicting localized and metastatic disease, respectively. The precisions were 0.80 and 0.68, the recalls were 0.66 and 0.88, and the F1 Scores were 0.72 and 0.77.

The machine learning (ML) model achieved accuracies of 0.95 and 0.86 for predicting localized and metastatic disease, respectively. The precisions were 0.95 and 0.78, the recalls were 0.99 and 1.00, and the F1-scores were 0.97 and 0.88.

The combined approach achieved accuracies of 0.96 and 0.88 for predicting localized and metastatic disease, respectively. The precisions were 0.95 and 0.81, the recalls were 1.00 and 1.00, and the F1-scores were 0.97 and 0.89.

## Discussion

In this study, the transformer-based machine learning (ML) model outperformed the baseline rule-based model for automatically classifying cancer trials with regard to the inclusion of patients with localized and or metastatic disease. However, allowing the rule-based model to overrule the predictions of the ML model as part of a combined approach resulted in an even better performance, even though the difference was small. This might be an indication that with a slightly larger training set, even better performance of the ML model could be achieved, as it should not be too difficult for the model to learn patterns in the title that can be formulated as fairly simple regular expressions. However, a model that makes its prediction based on the abstract of a publication can likely never achieve perfect performance as long as the information is not always contained in the abstract. Therefore, adhering to guidelines such as CONSORT that require information on eligibility criteria to be included in the abstract is important, not only for the model presented here, but for text mining approaches in evidence-based medicine in general.^12^ While it is a crucial piece of information for many cancer trials whether patients with localized and/or metastatic disease were eligible for inclusion, which should probably be part of the abstracts, the annotator had to open the full text of a publication several times to make an assessment during data labeling.

Notably, the combined approach had a perfect recall on the test set, i.e., it did not miss any trials that allowed for the inclusion of patients with localized and/or metastatic disease. This is arguably the behavior one would prefer in most contexts where a model like this could be deployed. E.g., when conducting a systematic review and using the model for screening trials that investigated therapeutic options for a particular tumor entity in the metastatic setting, it would be better to manually remove the trials that incorrectly make it through the screening due to the relatively lower precision than to have the model remove trials that could actually be included in the synthesis. However, the slight inclination to label a trial as allowing for the inclusion of patients with localized and/or metastatic disease could also be a disadvantage when trying to identify cancer screening or prevention trials.

This study has several limitations. First, we only used trials from seven journals for training and testing. While these are probably the journals that publish most practice-changing randomized controlled trials in oncology, we can’t assess the model’s ability to generalize to trials from other journals, especially those that use unstructured abstracts. In addition, the model is trained on oncology randomized-controlled trials, which means that these trials need to be identified first, either through automated measures or manually. However, at least for identifying randomized-controlled trials, several well-performing models have been described in addition to the fact that the CONSORT statement recommends explicitly identifying randomized controlled trials as such in the title.^13,14^ Lastly, confidence intervals were estimated using normal approximation intervals, while bootstrapping different training sets would likely have resulted in a more accurate estimate. However, training several hundred transformer models seemed excessive, considering the only marginal gain in information. To enable readers to judge the performance of the model on other cancer trials, a filter based on the model presented here can be tested on https://www.scantrials.com/.

The strengths of this study include the use of a dedicated unseen test set and the high degree of reproducibility as all code and annotated data are shared in a public repository. As an outlook, one can try to extract more inclusion as well as exclusion criteria and other characteristics from clinical trials by turning them into classification problems. While this approach has limitations, it seems feasible, at least for elements that are commonly mentioned in the title or abstract.

In conclusion, automatic classification of cancer trials with regard to the inclusion of patients with localized and or metastatic disease is feasible. Turning the extraction of trial criteria into classification problems could, in selected cases, improve text-mining approaches in evidence-based medicine.

## Data Availability

Data is currently available online at https://github.com/windisch-paul/metastatic_vs_local. Submission to a repository (Dryad) will be initiated after the submission of the preprint.

## References

1. Kilicoglu H, Rosemblat G, Hoang L, et al. Toward assessing clinical trial publications for reporting transparency. J Biomed Inform. 2021;116:103717.

2. Schmidt L, Sinyor M, Webb RT, et al. A narrative review of recent tools and innovations toward automating living systematic reviews and evidence syntheses. Z Evid Fortbild Qual Gesundhwes. 2023;181:65–75.

3. Saiz FS, Sanders C, Stevens R, et al. Artificial Intelligence Clinical Evidence Engine for Automatic Identification, Prioritization, and Extraction of Relevant Clinical Oncology Research. JCO Clin Cancer Inform. 2021;5:102–111.

4. Marshall IJ, Nye B, Kuiper J, et al. Trialstreamer: A living, automatically updated database of clinical trial reports. J Am Med Inform Assoc. 2020;27(12):1903–1912.

5. Yang Y, Jayaraj S, Ludmir E, Roberts K. Text Classification of Cancer Clinical Trial Eligibility Criteria. AMIA Annu Symp Proc. 2023;2023:1304–1313.

6. Greene FL, Sobin LH. The staging of cancer: a retrospective and prospective appraisal. CA Cancer J Clin. 2008;58(3):180–190.

7. Brierley JD, Gospodarowicz MK, Wittekind C. TNM Classification of Malignant Tumours. John Wiley & Sons; 2017.

8. Sheehan J, Hirschfeld S, Foster E, et al. Improving the value of clinical research through the use of Common Data Elements. Clin Trials. 2016;13(6):671–676.

9. Bao Y, Deng Z, Wang Y, et al. Using Machine Learning and Natural Language Processing to Review and Classify the Medical Literature on Cancer Susceptibility Genes. JCO Clin Cancer Inform. 2019;3:1–9.

10. Liu Y, Ott M, Goyal N, et al. RoBERTa: A Robustly Optimized BERT Pretraining Approach. arXiv [csCL]. Published online July 26, 2019. http://arxiv.org/abs/1907.11692

11. Kingma DP, Ba J. Adam: A Method for Stochastic Optimization. arXiv [csLG]. Published online December 22, 2014. http://arxiv.org/abs/1412.6980

12. Hopewell S, Clarke M, Moher D, et al. CONSORT for reporting randomised trials in journal and conference abstracts. Lancet. 2008;371(9609):281–283.

13. Marshall IJ, Noel-Storr A, Kuiper J, Thomas J, Wallace BC. Machine learning for identifying Randomized Controlled Trials: An evaluation and practitioner’s guide. Res Synth Methods. 2018;9(4):602–614.

14. Begg C, Cho M, Eastwood S, et al. Improving the quality of reporting of randomized controlled trials. The CONSORT statement. JAMA. 1996;276(8):637–639.

